# Communicating the health risks of wildfire smoke exposure: Health literacy considerations of public health campaigns

**DOI:** 10.1101/2020.09.14.20194662

**Authors:** Lily A. Cook

## Abstract

Effective communication about the health effects of wildfire smoke is important to protect the public, especially those most vulnerable to the effects of exposure: people with chronic respiratory conditions, children, and older adults. The objective of this paper is to examine the clarity and accessibility of materials intended to provide education about the health effects of wildfire. The Centers for Disease Control’s Clear Communications Index (CCI) is used to evaluate whether materials adhere to the main principles of health literacy: clarity and accessibility. This analysis found that only 32% of the materials received a passing score on the Clear Communications Index. Most materials were successful at clearly presenting specific behavioral recommendations, particularly that people should avoid exposure to air polluted by wildfire smoke by staying indoors, reducing activity levels, and using air purifiers or approved dust masks. However, materials often failed to acknowledge any uncertainty around these recommendations. Creators of these materials may want to incorporate more relevant illustrations to support the main message, and consider how information about the risks and benefits of the recommended behaviors can most clearly be presented.

## Introduction

Due to climate change, the intensity and frequency of wildfires has been increasing in the western United States (Westerling et al., 2006), causing growing numbers of people to be exposed to wildfire smoke and its subsequent health impacts. As a result, there is an emergent need to educate the public about the issue, and to provide materials that can be easily understood by the layperson. Because smoke exposure is episodic and only lasts between a couple of hours and several days, it is possible to prevent the worst effects by taking safeguards such as staying indoors. Educating the public allows them the opportunity to take steps to protect themselves from the health effects of wildfire smoke, the most well-documented of which is the exacerbation of pre-existing respiratory disease.

Educational public health messages are far more likely to have an impact if the precaution is communicated in a way that is engaging and accessible (Doak et al., 1996). The clarity of the messaging about the health effects of wildfire smoke, therefore, is an important public health issue. Despite this, there is a dearth of research formally evaluating these messages; in fact, a search of PubMed conducted in March of 2021 for the terms “‘health literacy’ AND ‘wildfire’” produced zero results. Additional searches were unable to find research addressing any of the following questions: How many informational materials about the health impact of wildfire smoke are available to the public? Who is producing them? How accessible are these materials to people with varying degrees of health literacy? What information do these materials present?

The objectives of this analysis were to (a) provide a descriptive overview of the materials available to the public about the health impact of wildfire smoke, and (b) analyze which components of the messaging are most accessible and which elements may need adjustment in order to be more clearly understood by the layperson.

## Materials and methods

### Data sources and searches

An internet search was conducted during February and March of 2019 to gather a comprehensive sample of publicly available materials about the physical health effects of wildfire smoke exposure. Most were collected using snowball sampling of resource lists provided by state and local government. Materials were gathered until saturation was reached; that is, until the author was unable to identify any new materials or resource lists. A list of inclusion and exclusion criteria is listed in Table 1.

**Table 1.**
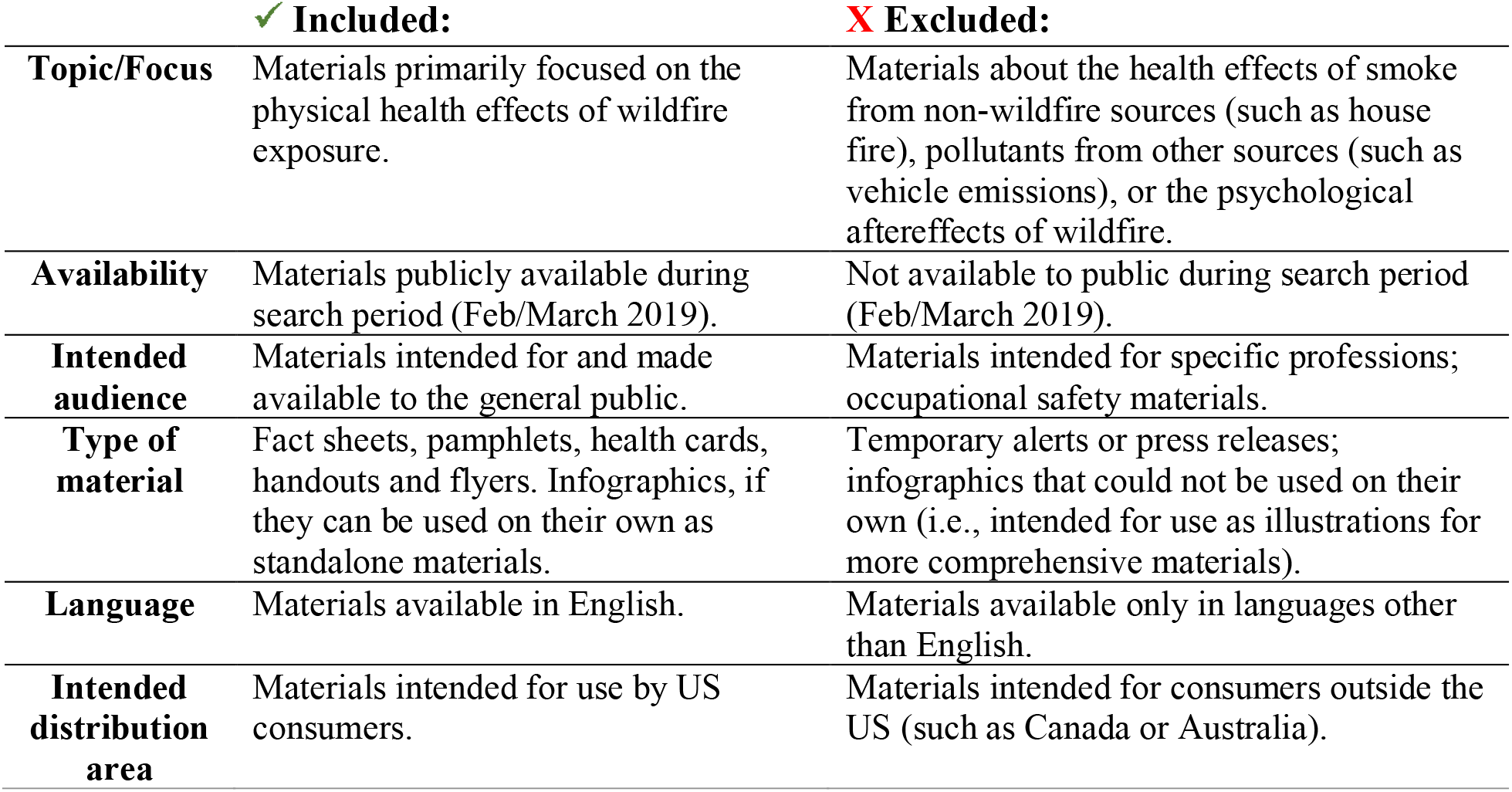
Inclusion and exclusion criteria

|  | ✓ **Included:** | ✗ **Excluded:** |
| --- | --- | --- |
| **Topic/Focus** | Materials primarily focused on the physical health effects of wildfire exposure. | Materials about the health effects of smoke from non-wildfire sources (such as house fire), pollutants from other sources (such as vehicle emissions), or the psychological aftereffects of wildfire. |
| **Availability** | Materials publicly available during search period (Feb/March 2019). | Not available to public during search period (Feb/March 2019). |
| **Intended audience** | Materials intended for and made available to the general public. | Materials intended for specific professions; occupational safety materials. |
| **Type of material** | Fact sheets, pamphlets, health cards, handouts and flyers. Infographics, if they can be used on their own as standalone materials. | Temporary alerts or press releases; infographics that could not be used on their own (i.e., intended for use as illustrations for more comprehensive materials). |
| **Language** | Materials available in English. | Materials available only in languages other than English. |
| **Intended distribution area** | Materials intended for use by US consumers. | Materials intended for consumers outside the US (such as Canada or Australia). |

### Evaluation framework

The Clear Communication Index (CCI) was used to rate the accessibility of the materials. The CCI was created in 2013 by the Center for Disease Control (CDC) in order to assess whether materials can be easily understood at all levels of health literacy understood (Baur & Prue, 2014). The Index emphasizes the importance of clarity when communicating health information, and was developed based on known best practices for public-facing materials, particularly the Suitability Assessment of Materials created by health literacy pioneers Cecilia and Leonard Doak (1996). The Clear Communication Index’s User Guide (2013) suggests four possible uses for the Index, including “to quickly assess the clarity and ease of use of an already released communication product.” Several studies have used the Clear Communication Index to evaluate the accessibility of materials intended to educate the public, including a study that looked at messaging in the drinking water reports provided by community water systems (Phetxumphou et al., 2016), an analysis of patient guidelines for common cancers (Tran et al., 2018), and one of patient materials for sickle cell disease (McClure et al., 2016).

The Clear Communication Index begins with four unscored, open-ended questions about the intended audience and message. Next, the Index asks yes or no questions which can be scored to create a rating of the overall accessibility of the material. Specific examples of criteria are provided on the score sheet in order to reduce ambiguity, and the score sheet also refers the evaluator back to the user guide for more comprehensive explanations if necessary. Table 2 includes a complete list of questions in the Index and their scoring criteria.

**Table 2.**
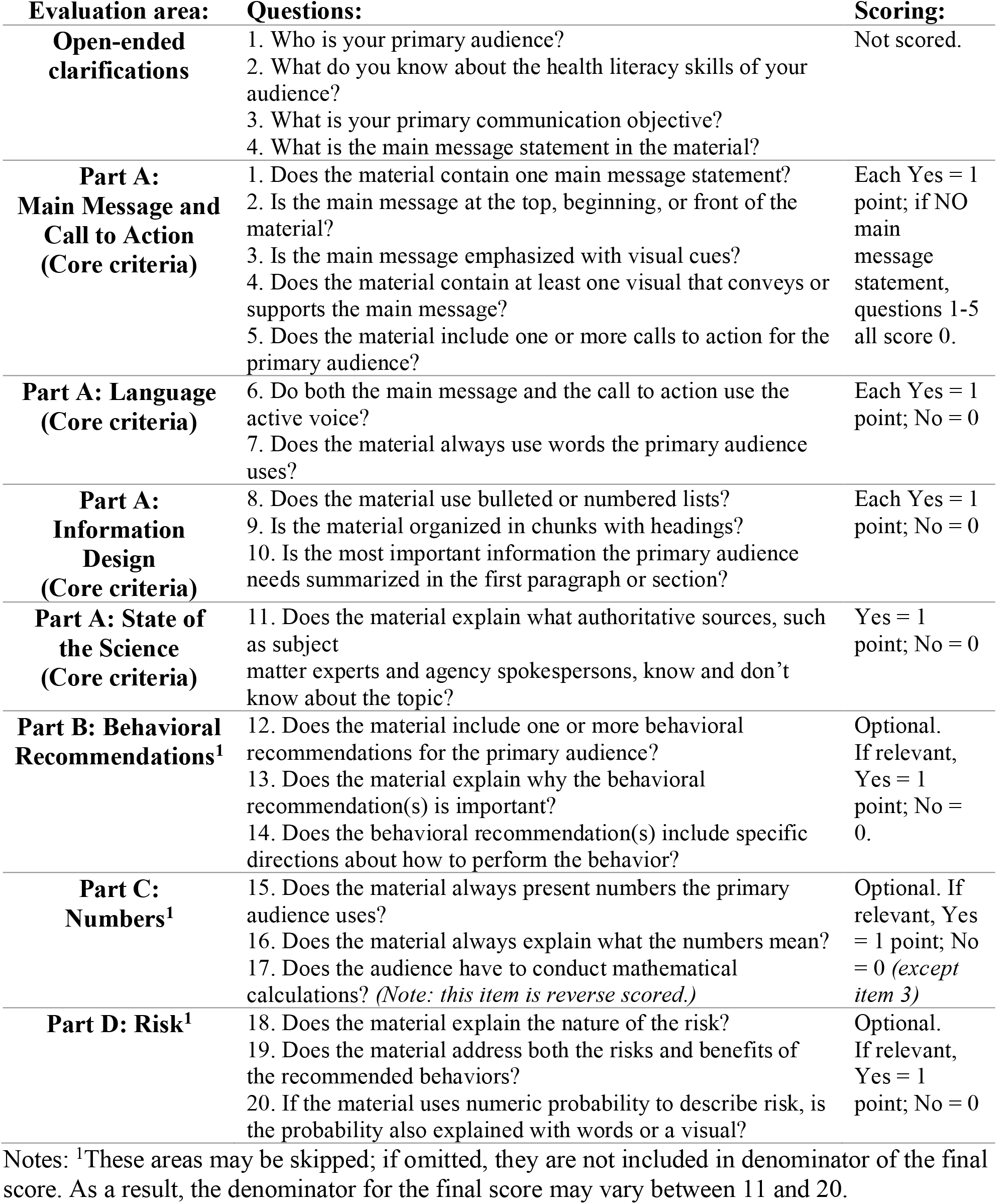
Evaluation areas and questions included in the Clear Communication Index (CDC, 2013)

The Index recommends a score of at least 90% for any given material to be considered accessible at all levels of health literacy. Although there are 20 questions in the full Clear Communication Index, it should be noted that the Index flexibly provides the ability to skip some areas if they do not apply to the material being evaluated; only the first 11 questions are considered core criteria and therefore always scored. This means that the overall number of points available may vary, because areas that do not apply are excluded from the denominator. To assure consistency, all materials evaluated for this study were scored and data extracted by a single rater. Descriptive statistics for the materials were calculated to learn more about which aspects of the health effects of wildfire smoke exposure are being most clearly communicated.

## Results

A total of 50 materials were located for evaluation; a full list of the titles of and sources for these materials can be found in Appendix A. Only 32% (n = 16) received a passing score of 90% or better on the Clear Communication Index; the mean score was 83% (SD 12%). Scores ranged from a low of 47% to a high of 100%. The length of the materials ranged from one page to five pages with a median length of one page.

### Core criteria

The average score on the core criteria (i.e., Part A) was 85% (SD 13%). Ninety-eight percent of the materials included a main message statement (Question 1); a typical main message was, “There are things you can do, indoors and out, to reduce your exposure to smoke” (US Environmental Protection Agency, n.d.-a). The core criterion materials were most likely to fail was the ‘State of the Science’ (Question 11). This criterion requires an acknowledgement of the uncertainty surrounding the central issue addressed in the material, which only 44% of materials (n = 22) did. Twenty-six of the materials (52%) neglected to include “at least one visual that conveys or supports the main message” (Question 4) (CDC Clear Communication Index: User guide, 2013). While most materials did include at least one graphic, oftentimes these graphics did not illustrate the main message; the User Guide to the CCI (2013) recommends that illustrations either (a) provide a caption or label contextualizing the illustration to the reader, or (b) demonstrate the behavior recommended in the material. By the criteria of the Clear Communication Index, therefore, most of these illustrations were inadequate.

### Behavioral recommendations, numbers, and risk

Although the ‘Behavioral Recommendations’ section (Part B) of the Clear Communication Index is optional, all the materials provided recommendations for specific behaviors and received a score for this section. The mean score for Part B was 84% (SD 18%). The most frequently recommended behaviors were to avoid exposure to smoky air by staying indoors, reducing activity levels, and using High Efficiency Particulate Air (HEPA) air purifiers or approved masks. The lowest scoring item in Part B concerned whether or not the material explained the importance of the behavior(s) to the audience (Question 13); only 62% (n = 31) of materials did so.

Only seven materials (14%) presented numerical information; as a result, the optional Part C (‘Numbers’) was the most likely to be omitted from the scoring. The small sample made it difficult to draw reliable conclusions about the manner in which numbers about wildfire smoke and human health are presented to the public.

The risks of wildfire smoke exposure were only addressed in 21 of the materials (42%), and of the four sections of the Clear Communication Index, the materials collectively received the lowest score for Part D, ‘Risk’ (58%). Of those that did address risk, 57% (12 out of 21) failed optional Question 19, which asks if both the risks and benefits of the recommended behaviors have been discussed in the material. For example, few materials pointed out that staying indoors with the windows closed, a commonly recommended behavior, may create a heat risk for those who live in hot areas and do not have air conditioning.

### Material creators

The most prolific producer of these materials is the US Environmental Protection Agency (US EPA), which created eight distinct publications (16%). The US EPA also collaborated with the US Forest Service, the Centers for Disease Control and Prevention, and the California Air Resources board to produce “Wildfire Smoke: A Guide for Public Health Officials,” a publication which was not assessed here but served as the basis for many of the materials (Lipsett et al., 2016). Seven states have produced materials tailored to the specific needs of their populations; six of the seven are located in the Western United States (California, Washington, Oregon, Idaho, Montana, and Arizona). At the local level, numerous air districts within California have also produced public health campaigns on wildfire smoke and human health. Seven of the materials (14%) were made available in languages other than English; five of these (71%) were from the Washington State Department of Health (2018), which has translated their educational materials into 10 languages other than English.

### Vulnerable populations

Some groups, such as children and the elderly, are considered to be more susceptible to the health effects of wildfire smoke, and seventeen materials (34%) were devoted primarily to the needs of these more vulnerable individuals. The mean Clear Communication Index score for the materials devoted to vulnerable groups was not significantly different from those intended for the general population (mean score 84% vs. 82%, p-value = 0.48). Six of these materials were intended for caregivers of children, and provided messages such as “there are things you can do to protect children’s health from the harmful pollutants filling the air due to wildfire smoke” (Air Districts of the Sacramento Region, n.d.). Five were intended to guide decisions about the safety of outdoor activities for children during smoke events, and four were specific to seniors. Also included in the review was a handout from the Oregon Veterinary Medical Association (2017) called “Wildfire Smoke & Animals.” This pamphlet is intended to educate the public on how wildfire smoke can affect an often-overlooked group: pets. The main message of that material is that “health advisories for air quality also apply to animals,” and it notes that pet birds are particularly susceptible to the effects of wildfire smoke.

## Discussion

There was overlap in materials produced at the state and local level, which meant that this review was able to compare the communication strategies from different organizations promoting a similar health message. For example, although four states (Montana, Washington, Oregon, and California) presented versions of the same message about air pollution and outdoor activities, Washington’s material scored 92% while Montana’s received the lowest score of all the evaluated materials: 47%. While Washington’s material, “Air Pollution and School Activities” (2015), explicitly states the single main message (“Exercising students breathe deeper and more often and take in more air, and more air pollution, into their lungs. Breathing polluted air can cause health problems, including aggravating asthma and other respiratory diseases”), Montana’s version scored poorly because it completely omitted the main message, and thus failed to make it clear that limiting activity during smoky periods can help reduce the health effects of wildfire smoke exposure (Montana Department of Environmental Quality, 2016). Montana’s material also received a lower score due to its use of overly complex and ill-defined language.

As noted in the results, few of the materials reviewed here explained why it is important to protect oneself from wildfire smoke exposure. It is possible, however, that this message was omitted because it is assumed that the risks of breathing wildfire smoke are obvious; historians have noted that the health risks of air pollution seem to be intuitively understood, as evidenced by the fact that people have demonstrated concern about the health effects of air pollution as far back as the 17th century (Lipfert, 1994).

The finding that these materials often neglect to provide information about both risks and benefits for the recommended behaviors might be the result of an attempt by material authors to avoid confusing the public with contradictory messages. For example, during 2018’s Camp Fire in California, the city of Sacramento handed out N95 masks to residents at the same time that Sacramento County warned that these masks can “lead to breathing difficulties and elevated heart rate, especially for people with respiratory or cardiac conditions” (“Are N95 smoke masks safe? One California county recommends against use,” 2018). Local media reported that this warning confused some members of the public by created the impression that those with respiratory and cardiac conditions should not use the masks, although they are in fact the group most in need of them.

Finally, it should be noted that there is a necessary trade-off between length of a public health message and its simplicity. Because disease often has complex pathophysiological mechanisms, some complexity must be omitted in order to create a message simple and brief enough to be easily understood by people at all levels of health literacy. In order to keep the message accessible, those who create materials for a general audience must walk a precarious tightrope between simplicity and complexity, careful not to sacrifice too much of either. With materials about the health effects of wildfire, creators appeared to optimize this balance in two different ways:

1. *With a series of materials*. At the federal level, the US EPA produced a series of five Wildfire Smoke Factsheets intended to increase preparedness among people living in wild-fire-prone areas (US Environmental Protection Agency, n.d.-b). Topics include selection and proper use of a face mask, tips for protecting children from the health effects of wildfire smoke, selecting a HEPA filter, etc. At the state level, Washington state’s Smoke from Wildfires Toolkit addresses a variety of topics over several fact sheets (Washington State Department of Health, 2018).
2. *With the use of graphics*. When appropriately used, graphics can present information visually in a way that can summarize even complex recommendations simply and succinctly (Houts et al., 2006).

### Future directions

Several materials mentioned that children should not use adult-sized N95 masks during wildfire smoke events, but did not provide any alternatives or specify why. Are child-sized N95 masks available, or even advisable? Is it better for children to use an adult mask than no mask at all? Hopefully future research will inform the next generation of public health messaging, which will be able to provide clearer advice about the issue.

Although public health campaigns have traditionally disseminated information as pamphlets or fact sheets, the digital revolution has made it easier to create interactive materials. Future public health campaigns may go beyond merely providing information, and have the capability to interact with the public via apps or mobile technologies. At the time of this writing, there is only one interactive material intended to educate the public about the health effects of wildfire smoke: Smoke Sense. Created by the US EPA (n.d.-c), Smoke Sense functions as a crowd-sourcing app, provides education about air quality, and also delivers current and forecasted smoke conditions. Although Smoke Sense is intended primarily as a citizen science app, its creators have been analyzing user-generated data and using these insights to better understand how to raise public awareness about the health risks of wildfire smoke (Rappold et al., 2019).

Finally, little is known about whether these messages effectively protect public health. Because the issue has only recently acquired public prominence, these campaigns are relatively new, and these messages may need more time to disseminate before their effectiveness can be judged.

## Conclusions

A variety of materials designed to educate the public about how to protect their health from wildfire smoke are currently available, some catered to the needs of vulnerable populations and available in multiple languages. While these materials do a good job of providing specific behavioral recommendations about how to keep safe from wildfire smoke, most did not receive a passing grade on the Clear Communication Index, usually because they either failed to provide relevant visual information or they did not address the current state of the science. Future iterations may benefit from incorporating illustrations to support their message and updated information about the risks and benefits of the recommended behaviors from experts on wildfire and human health. Additional research about uptake is necessary to determine if these messages have successfully protected the public from the health effects of wildfire smoke exposure.

## Data Availability

Web links to the materials referred to in the manuscript can be found in the appendix. Web links are all active at the time of this writing, but are also available from the author upon request.

## Acknowledgments

Thank you to Dr. Nicole Weiskopf for providing helpful feedback on this manuscript.

## Funding

This work is supported by National Library of Medicine and the National Institute of Environmental Health Sciences under Award #T15LM007088. The content is solely the responsibility of the author and does not necessarily represent the official views of the grantor(s).

## Institutional Review Board Statement

Not applicable.

## Informed Consent Statement

Not applicable.

## Data Availability Statement

Appendix A contains a list of all the materials assessed in this article, links to their locations, and other information that may be useful for validating or reproducing the results found here. Requests for additional data not found in the Appendix can be submitted to the corresponding author.

## Conflicts of Interest

The author declares no conflict of interest. The funders had no role in the design of the study; in the collection, analyses, or interpretation of data; in the writing of the manuscript, or in the decision to publish the results.

## Appendix A: Materials assessed

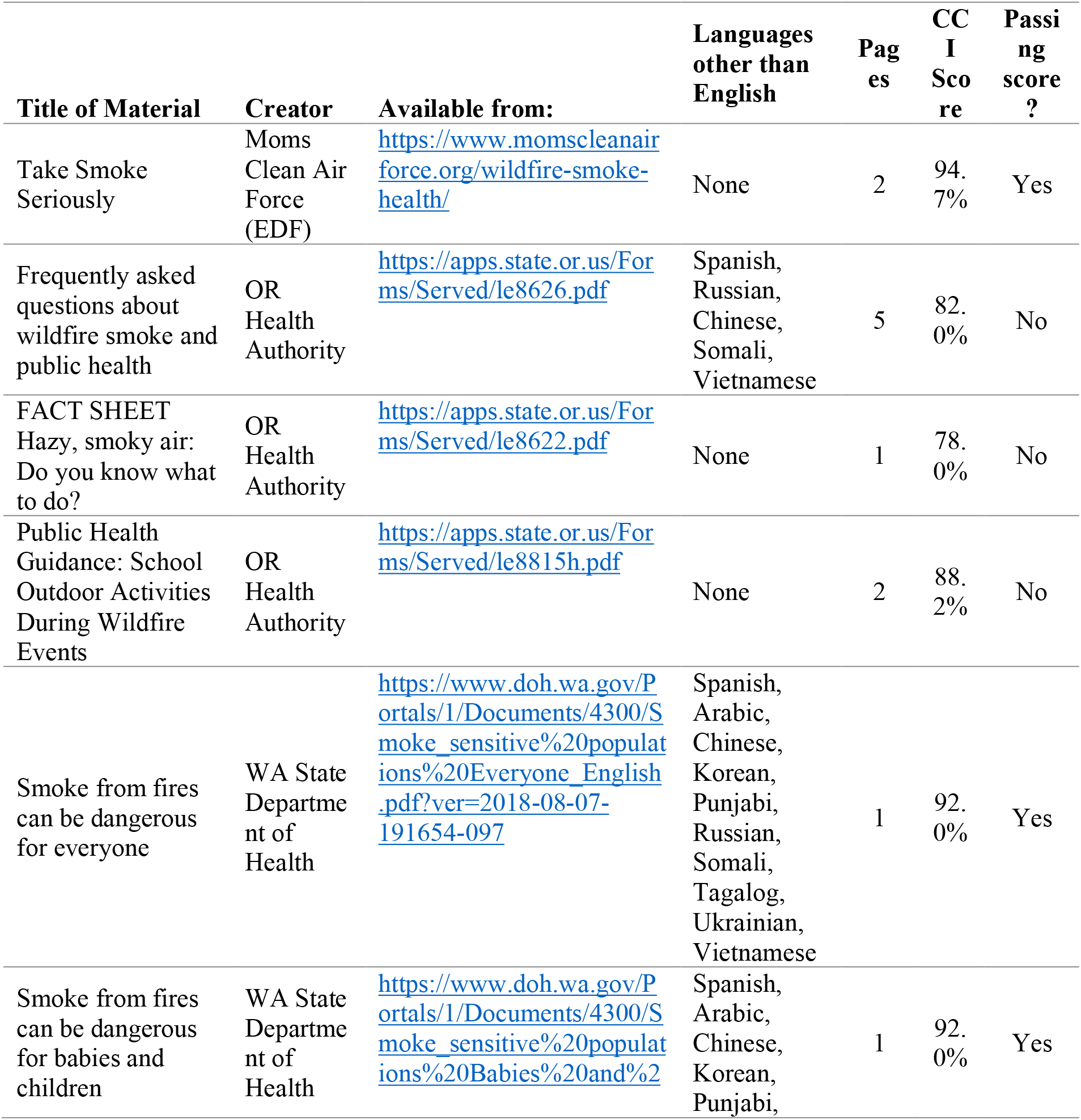
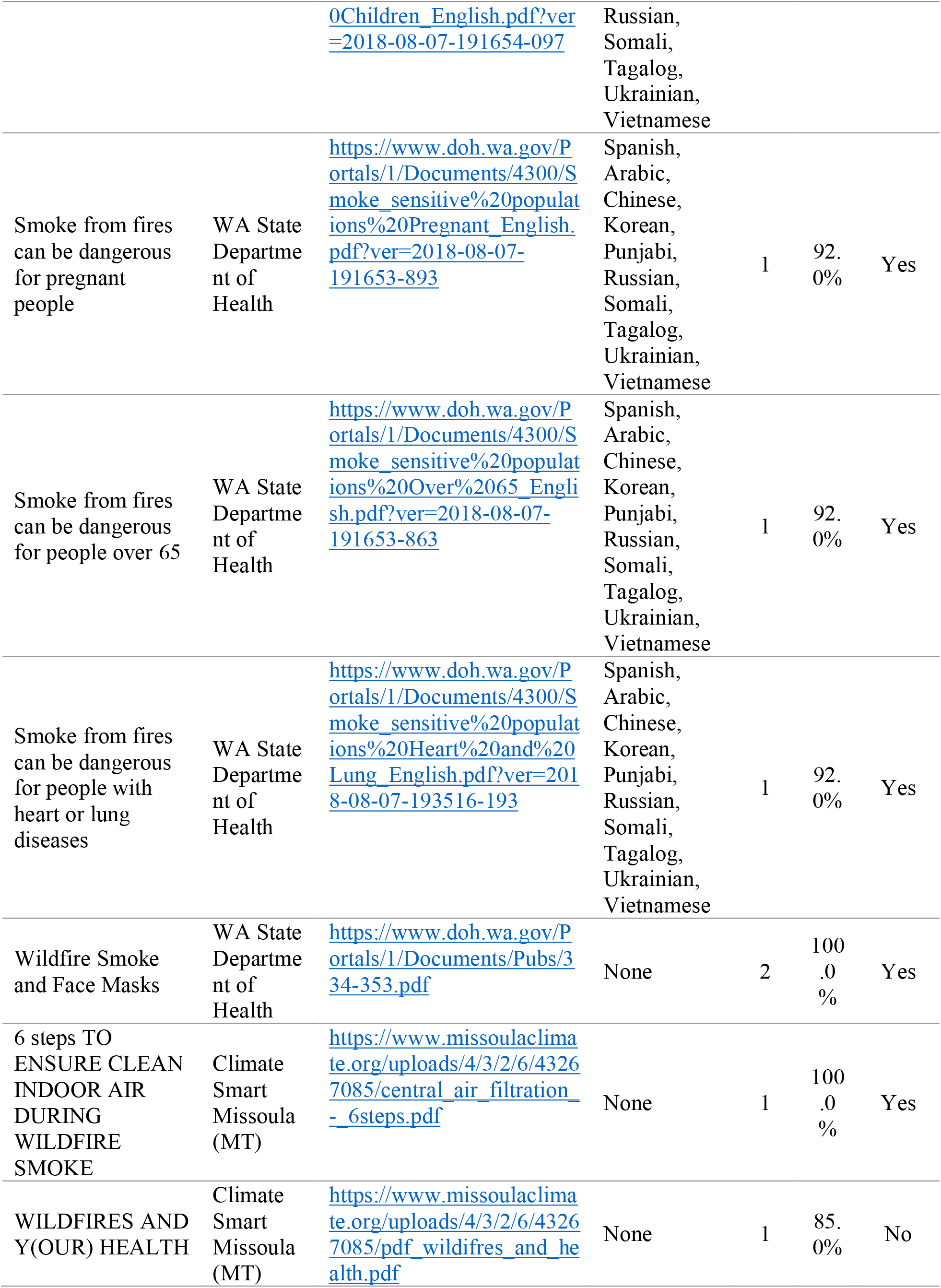
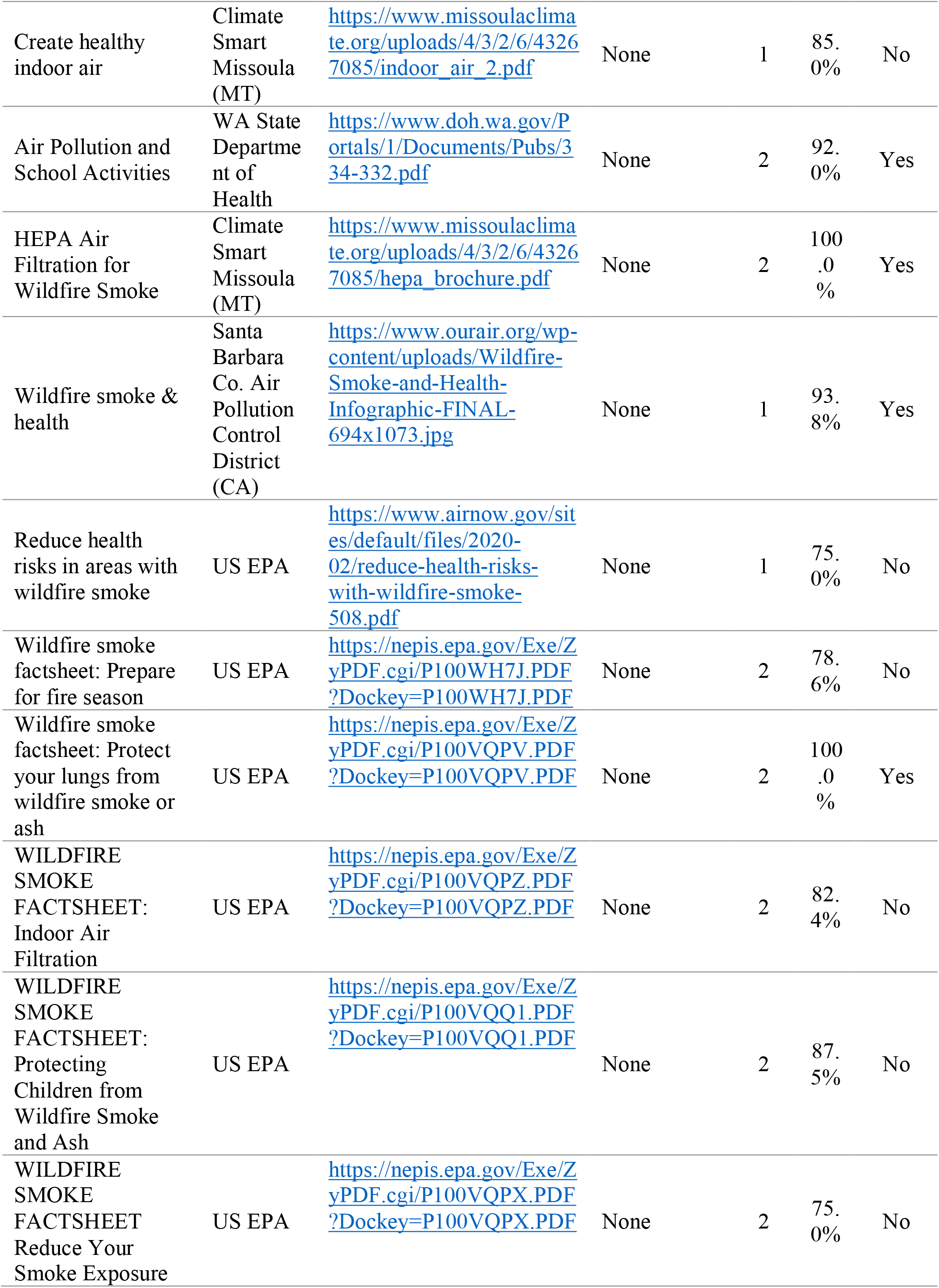
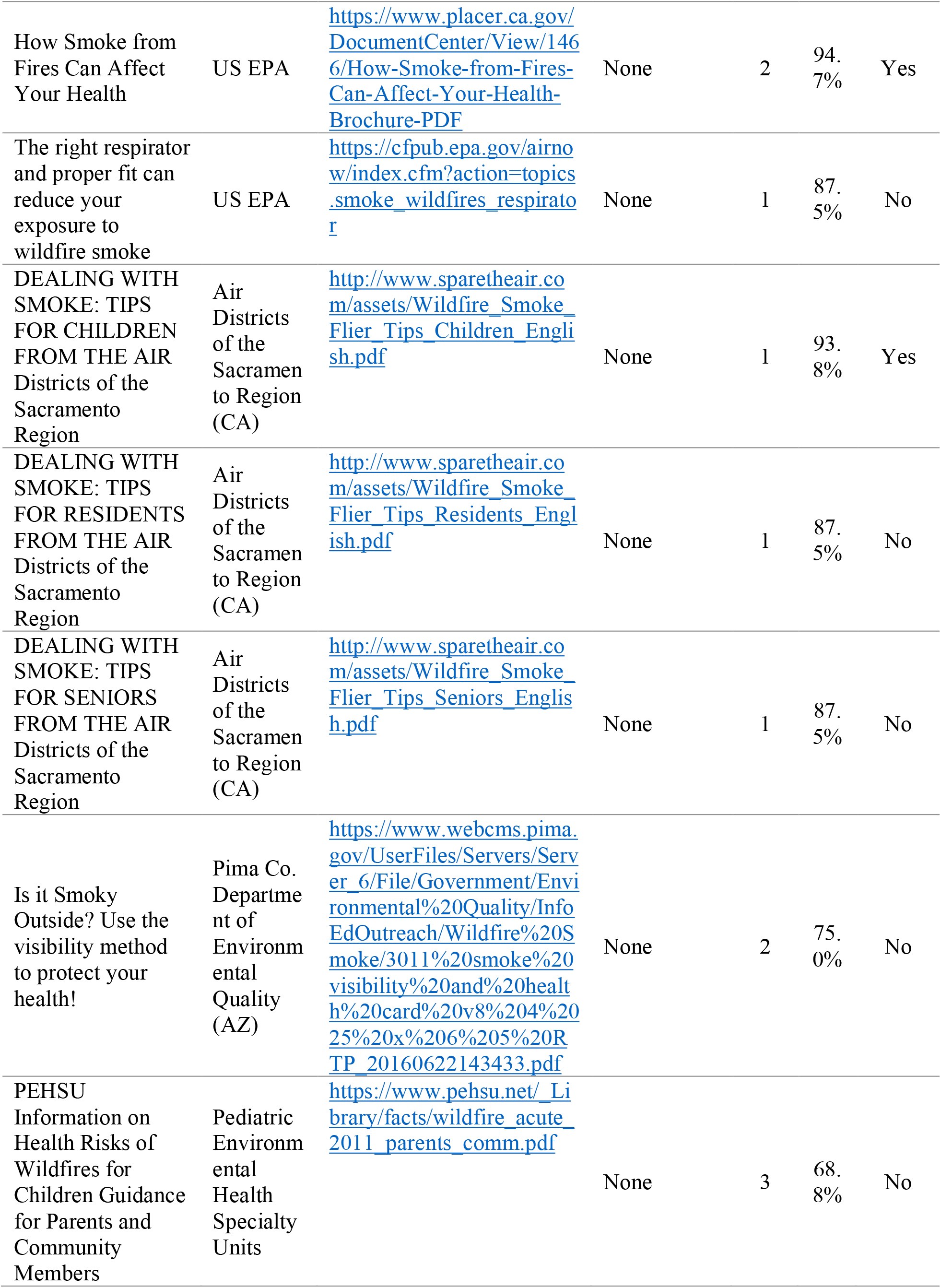
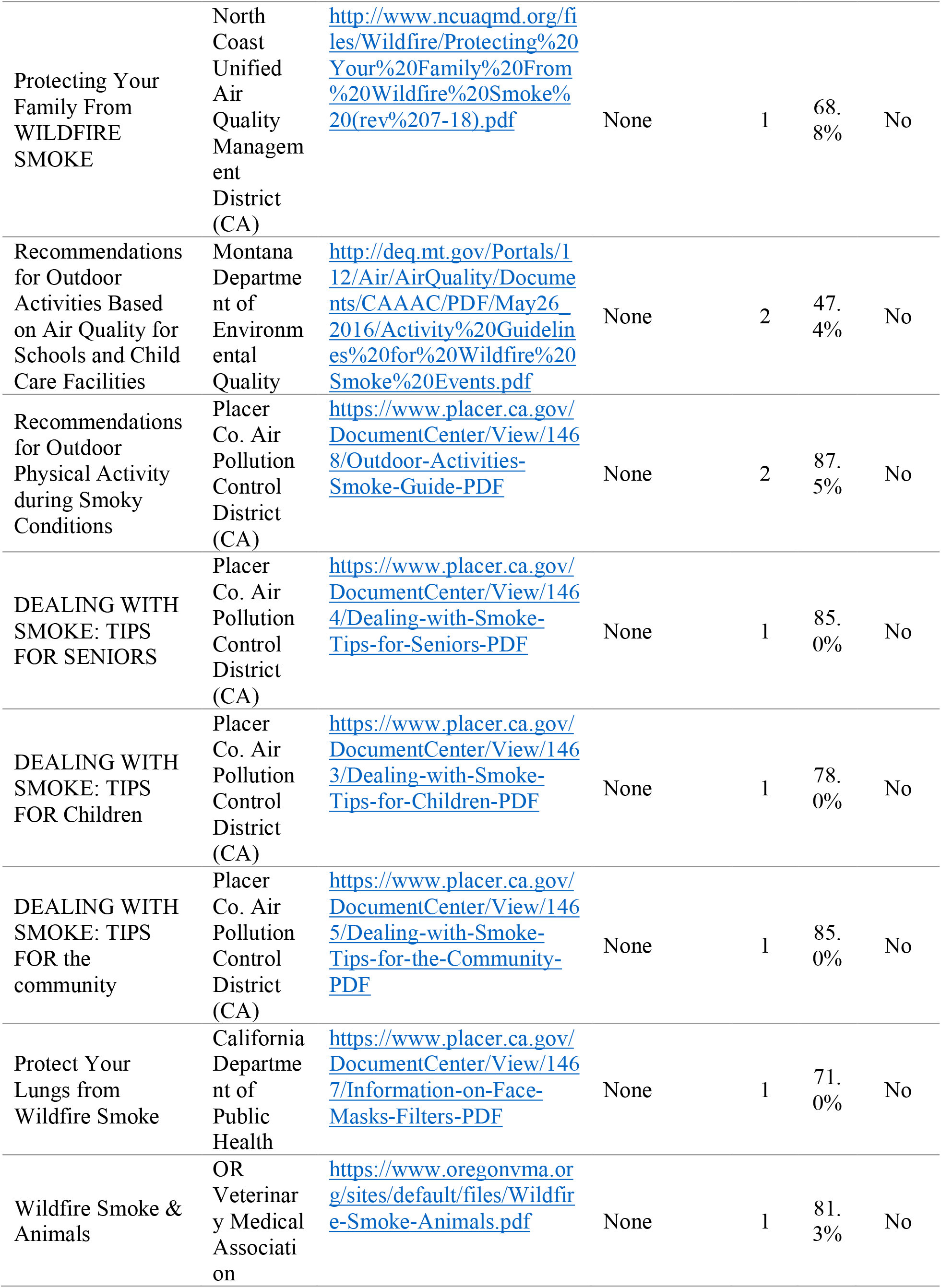
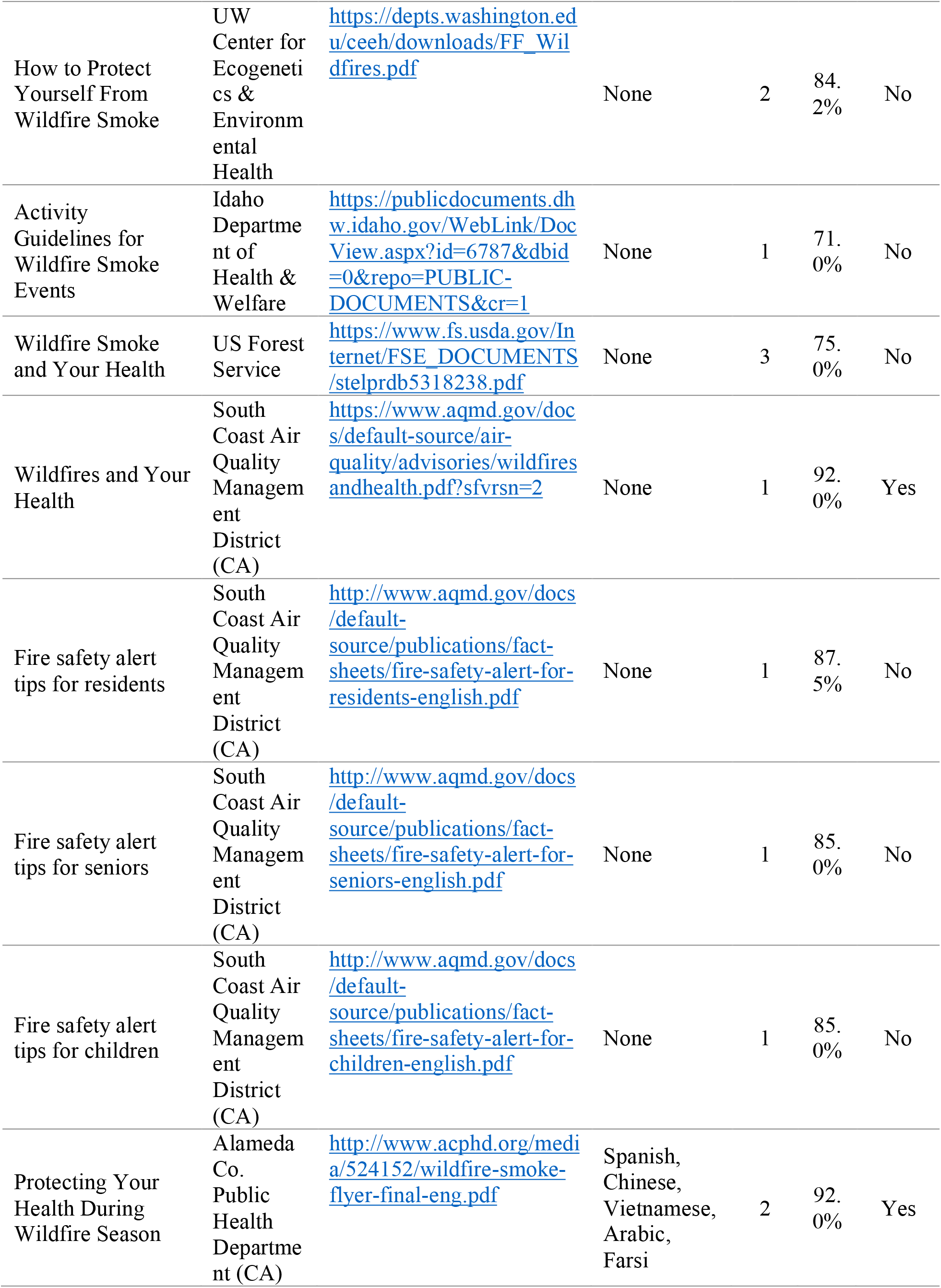
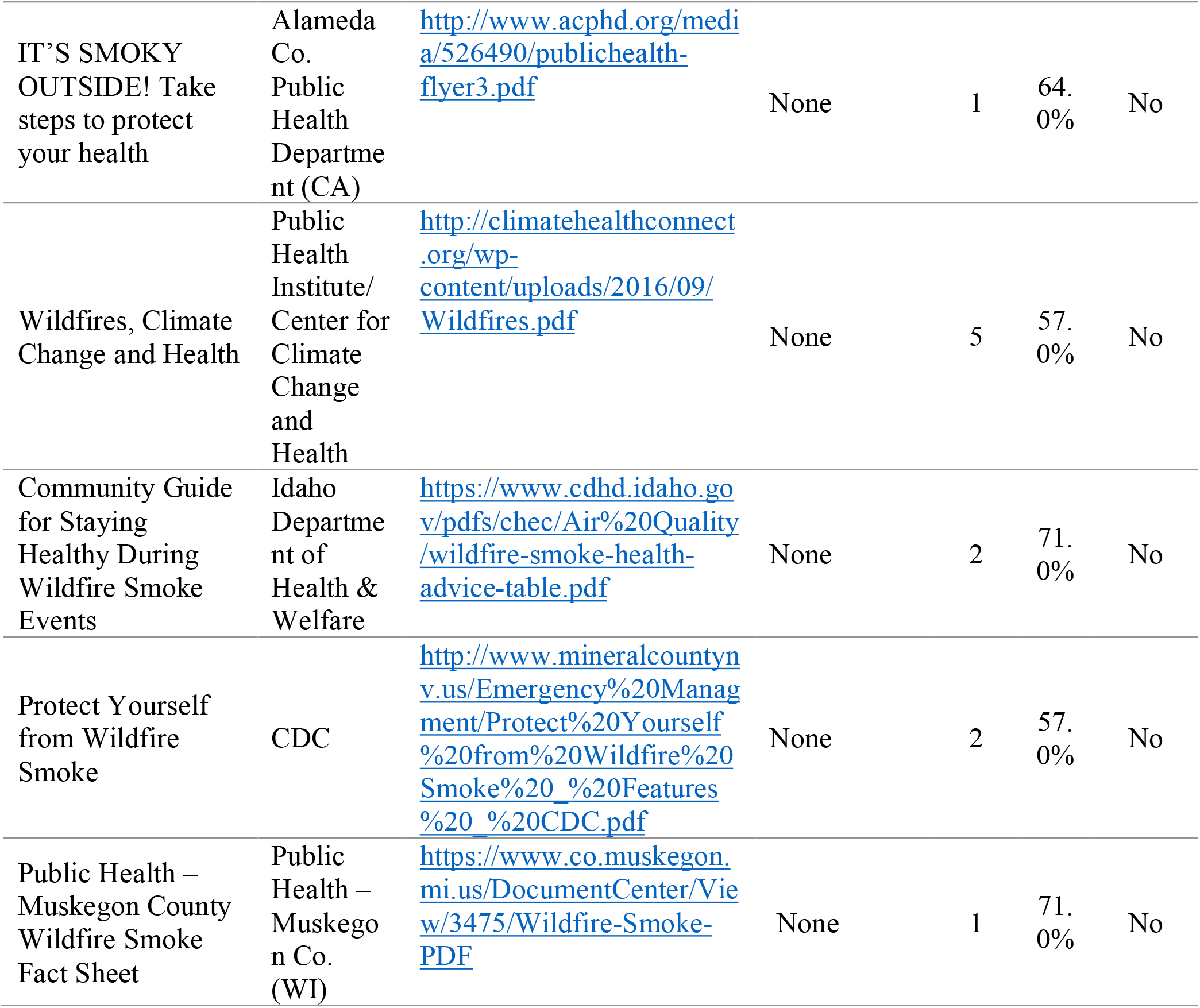

## Notes

### Competing Interest Statement

The authors have declared no competing interest.

### Funding Statement

This research supported by the National Library of Medicine and the National Institute of Environmental Health Sciences training grant, under Award Number T15LM007088.

### Author Declarations

No IRB approval was necessary for this work, as it is a review of public health campaigns.

### Summary of Updates

More detailed case analysis. Updated for clarity.

